# Association of Kras mutation with tumor deposit status and overall survival of colorectal cancer

**DOI:** 10.1101/19003210

**Authors:** Meifang Zhang, Wenwei Hu, Kun Hu, Yong Lin, Zhaohui Feng, Jing-Ping Yun, Nan Gao, Lanjing Zhang

## Abstract

**Background:** The recent staging manual upstages Node-negative tumor-deposit positive colorectal cancer (CRC) from N0 to N1c category, while the development of tumor-deposit presence is poorly understood. Meanwhile, Kras mutation is associated with progression of CRC, but its link to tumor-deposit status is unclear.

**Method:** This retrospective cohort study included the patients with incidental CRC diagnosed during 2010-2014 in the National Cancer Database and recorded statuses of Kras and tumor deposit. We conducted multivariable logistic regression and Cox regression analyses to investigate the factors associated with tumor-deposit status and overall-survival, respectively.

**Results:** A total of 48,200 CRC patients with Kras status were included in the study (25,407 [52.7%] men, 25,648[46.8%] <65 years old, 18 381 [38.1%] with Kras mutation). Adjusted for microsatellite instability, age, pathologic stage and tumor grade, Kras mutation (versus wild-type) was associated with tumor-deposit presence (n=15,229, odds ratio=1.11, 95% CI 1.02 to 1.20). Kras mutation was also independently linked to a worse overall survival of CRC patients regardless of tumor-deposit status (n=8,110, adjusted Hazard ratio=1.40, 95% CI 1.09 to 1.79 for CRC with tumor deposits, and n=2,618, adjusted HR=1.63, 95% CI 1.16 to 2.28 for CRC without), but a better survival in CRC with no known/applicable tumor-deposit status (n=457, adjusted Hazard ratio =0.32, 95% CI 0.11 to 0.95).

**Conclusion:** Kras mutation is independently associated with tumor-deposit presence, and a worse overall survival of CRC with or without tumor-deposit. Therefore, it may play a role in the development of tumor deposits and serve as a target for CRC treatment.

## Introduction

Colorectal cancer (CRC) is the third most common cancer and a leading cause of cancer death in the United States.[1] Kirsten rat sarcoma viral oncogene homolog (Kras) is a proto-oncogene that plays an important role in the development and treatment of CRC. Its mutation occurs in approximately 30 to 45% of CRC, and mostly in codon 12 or 13.[2-7] The patients with Kras mutation are unlikely to benefit from anti-EGFR (epidermal growth factor receptor) therapy, which should thus be applied to only the CRC with wild type Kras as recommended by the National Comprehensive Cancer Network (NCCN) guidelines.[8] However, the association between Kras mutation and patients’ survival remained controversial. A clinical trial showed that a Kras mutation was a strong negative prognostic factor,[9] while other reports failed to show any prognostic value of Kras.[10, 11] Nonetheless, the NCCN guidelines for Treatment of Colon/Rectal Cancer suggested that all patients with metastatic CRC should be treated with the detection of Kras mutations.[8]

The American Joint Committee on Cancer (AJCC) tumor node metastasis (TNM) staging system is widely used to predict prognoses in CRC patients and to guide adjuvant therapy. The 7^th^ and 8th editions of the manual (AJCC 7; 2010; AJCC 2018) classified extranodal tumor deposits (TDs) that lack regional lymph node metastasis as N1c in TNM staging system.[12, 13] The presence of TDs was associated with a poorer outcome as illustrated by decreased disease-free survival (DFS) and overall survival (OS).[14, 15] TDs-positive CRC patients have been proposed to be treated as stage III and have been shown to be strongly associated with worse DFS, compared with higher N stages.[16] However, our recent work also showed the upstaging node-negative CRC with TD (N0 to N1c) led to more chemotherapy and 43% more all-cause mortality.[17] Therefore, it is not clear how to best classify and treat CRC with TD. The associations of Kras status with TD status and overall survival of CRC are also unclear.

The purpose of the retrospective cohort study was to investigate the association of Kras mutation with TD status and overall survival of CRC using a large cancer database.

## Methods

The National Cancer Database (NCDB), established in 1989, is a nationwide, facility-based, comprehensive clinical surveillance resource oncology data set. It is the largest clinical cancer registry in the world and a joint project of the American Cancer Society and the Commission on Cancer of the American College of Surgeons.[18] The NCDB extracts data through all available components of the medical record by certified tumor registrars at all cancer centers accredited by the American College of Surgeon’s Commission on Cancer. A rigorous program has been implemented to ensure data quality. Scholars have used NCDB to investigate colorectal, breast and lung cancers.[19-23] An Institutional Review Board (IRB) review was exempt for this study due to the use of publicly available, deidentified, existing database (Exempt category 4).

In this retrospective cohort study on the 2017-release of NCDB (followed through Dec. 2014), the primary end point was the tumor-deposit status, which was dichotomized using the data of site-specific factor 4. The secondary endpoint was the overall survival. No cancer specific survivals could be assessed due to the lack of data on the death cause in the NCDB.[18] The inclusion criteria were all incident CRC cases diagnosed during 2010-2014, with data of Kras status, which became part of the NCDB (as site-specific factor 9) for CRC in 2010. The exclusion criterion was no surgical resection of primary tumor (code 998 for site-specific factor 4). We included the following factors in the univariate regression analyses: age, sex, tumor location (colon versus rectum), MSI status, Kras status, pathologic tumor stage (the 7^th^ AJCC staging manual, according to the data item TNM_EDITION_NUMBER), tumor grade (high versus low), race, Charlson-Deyo score, chemotherapy status, and radiotherapy status. A factor will be included in multivariable regression analyses if its *P* in the univariate analysis was less than 0.10. Specifically, MSI status was classified as stable/low (codes 20 and 40) and unstable/high (codes 50 and 60) because of its potential predictive value for chemotherapy outcome.[24] Races were classified as non-Hispanic (NH) White, NH Black, Hispanic and Others according to the data items of race and Hispanic ethnicity. Chemotherapy statuses were classified as received if chemotherapy data item was coded as chemotherapy (not otherwise specified) administered, single-agent chemotherapy or multiagent chemotherapy administered as first course therapy (codes 1, 2 and 3); otherwise as classified not received (codes 0, 86 and 87). Radiotherapy statuses were classified as received if a radiotherapy was indicated (codes 1-5), and classified as not received if no radiotherapy indicated (code 0).

We conducted statistical analyses using Stata (version 15, StataCorp LLC, College Station, TX). The Pearson’s Chi-square test and logistic regression models were used to assess potential associations. Multivariable Cox regression models with time-varying covariates were used for survival analyses, including the factors that had a *P* value less than 0.10 in univariate Cox regression models. All *P* values were two-sides, with *P*<0.05 as statistically significant.

## Results

Among the 514,964 incident CRC cases in the NCDB diagnosed during 2010-2014, 23,265 (4.52%) harbored a Kras mutation and 36,327 (7.05%) was wild type. A total of 455,372 (88.42%) cases had no known Kras statuses and were excluded. Among the 59,592 cases with known Kras status, 11,392 (19.1%) had no surgical resections of the CRC, and were excluded because the pathologic assessment of their tumor deposit status was not possible. The Chi-square test shows that all of included factors were associated with tumor-deposit status (**Table 1**), including age, sex, tumor location, MSI status, Kras status, pathologic tumor stage, tumor grade, race, Charlson-Deyo score, chemotherapy status, and radiotherapy status.

**Table 1.** Baseline characteristics of incident colorectal cancers with known Kras status in National Cancer Database diagnosed during 2010-2014

|  | Tumor deposit status |  |  |  |  |  | Total* | P |
| --- | --- | --- | --- | --- | --- | --- | --- | --- |
|  | None (%) |  | Yes (%) |  | NA (%) |  |  |  |
| **Total** | 31,568 |  | 11,814 |  | 4,818 |  | 48,200 |  |
| **Sex** |  |  |  |  |  |  |  | <0.001 |
| Male | 16552 | (52) | 6167 | (52) | 2688 | (56) | 25407 |  |
| Female | 15016 | (48) | 5647 | (48) | 2130 | (44) | 22793 |  |
| **Age** |  |  |  |  |  |  |  | <0.001 |
| <65 year | 16068 | (51) | 6806 | (58) | 2774 | (58) | 25648 |  |
| 65+ year | 15500 | (49) | 5008 | (42) | 2044 | (42) | 22552 |  |
| **Kras** |  |  |  |  |  |  |  | <0.001 |
| Wild type | 19775 | (63) | 7189 | (61) | 2855 | (59) | 29819 |  |
| Mutated | 11793 | (37) | 4625 | (39) | 1963 | (41) | 18381 |  |
| **Microsatellite Instability (MSI)** |  |  |  |  |  |  |  | <0.001 |
| Stable/Low | 10484 | (82) | 3605 | (85) | 698 | (84) | 14787 |  |
| Unstable/High | 2281 | (18) | 631 | (15) | 132 | (16) | 3044 |  |
| **Location** |  |  |  |  |  |  |  | <0.001 |
| colon | 27365 | (87) | 10649 | (90) | 3914 | (81) | 41928 |  |
| rectum | 4203 | (13) | 1165 | (10) | 904 | (19) | 6272 |  |
| **Tumor stage** |  |  |  |  |  |  |  | <0.001 |
| I-II | 12162 | (42) | 505 | (5) | 658 | (21) | 13325 |  |
| III-IV | 16980 | (58) | 10708 | (95) | 2490 | (79) | 30178 |  |
| **Grade** |  |  |  |  |  |  |  | <0.001 |
| Low | 23496 | (78) | 7256 | (64) | 2799 | (74) | 33551 |  |
| High | 6527 | (22) | 4059 | (36) | 1007 | (26) | 11593 |  |
| **Charlson-Deyo Score** |  |  |  |  |  |  |  | <0.001 |
| 0 | 22858 | (72) | 8908 | (75) | 3741 | (78) | 35507 |  |
| 1 | 6536 | (21) | 2226 | (19) | 815 | (17) | 9577 |  |
| 2 | 2174 | (7) | 680 | (6) | 262 | (5) | 3116 |  |
| **Race** |  |  |  |  |  |  |  | <0.001 |
| NH White | 23541 | (77) | 8840 | (77) | 3315 | (73) | 35696 |  |
| NH Black | 3656 | (12) | 1370 | (12) | 675 | (15) | 5701 |  |
| Hispanic | 2075 | (7) | 780 | (7) | 354 | (8) | 3209 |  |
| Others | 1322 | (4) | 496 | (4) | 228 | (5) | 2046 |  |
| **Received chemotherapy** |  |  |  |  |  |  |  | <0.001 |
| No | 9865 | (33) | 1473 | (13) | 864 | (19) | 12202 |  |
| Yes | 19613 | (67) | 9512 | (87) | 3665 | (81) | 32790 |  |
| **Received radiotherapy** |  |  |  |  |  |  |  | <0.001 |
| No | 27208 | (87) | 10470 | (89) | 4000 | (85) | 41678 |  |
| Yes | 4096 | (13) | 1247 | (11) | 691 | (15) | 6034 |  |

MSI, microsatellite instability; NH, Non-Hispanic; P, Chi-square test for associations; NA, not available; \*, Sum of the subtotals may not be equal to the grand total due to missing data in some strata.

Our multivariable logistic regression analysis showed that MSI (Unstable/High vs Stable/Low), Kras (Mutated vs Wild type), Age (65+ vs <65 year), Pathologic stage (III-IV vs I-II) and Tumor grade (High vs Low) were associated with presence of tumor deposit (versus absence), but not Charlson-Deyo Score, tumor location or sex (**Table 2**). Univariate Cox regression analysis and log-rank test both show that Kras mutation (versus wild-type) was linked to a worse overall survival of the CRCs, but no association between MSI status and overall survival (**Figure**). The multivariate Cox regression analyses also showed that the prognostic values of MSI and Kras statuses significantly differed by tumor deposit status (**Table 3**). In the CRC with or without tumor-deposits, Kras mutation (versus Wild type) was independently linked to a worse overall survival (*P*=0.008 for with tumor deposits, and *P*=0.004 for without); However, in the CRC with insufficient or no applicable data of tumor deposit status, Kras mutation (versus Wild type) was linked to a better overall survival (*P*=0.039, **Table 3**).

**Table 2.** Factors associated with tumor deposit status (present vs absent) of incident colorectal cancers in National Cancer Database diagnosed during 2010-2014 (n=15,229)

|  | OR | 95% CI | P |
| --- | --- | --- | --- |
| MSI (Unstable/High vs Stable/Low) | 0.84 | (0.75 to 0.95) | 0.004 |
| Kras (Mutated vs Wild type) | 1.11 | (1.02 to 1.20) | 0.015 |
| Location (Rectal vs Colonic) | 1.14 | (0.99 to 1.30) | 0.064 |
| Age (65+ vs <65 year) | 0.85 | (0.78 to 0.92) | <0.001 |
| Pathologic stage (III-IV vs I-II) | 14.44 | (12.51 to 16.66) | <0.001 |
| Tumor grade (High vs Low) | 1.58 | (1.45 to 1.73) | <0.001 |
| Charlson-Deyo Score | 0.94 | (0.87 to 1.01) | 0.081 |
| Race | 0.97 | (0.92 to 1.02) | 0.184 |
| Sex (Male vs female) | 0.96 | (0.89 to 1.04) | 0.338 |

P of multivariable logistic regression analysis; MSI, microsatellite instability; Race including Non-Hispanic White, Non-Hispanic Black, Hispanics and Others; OR, odds ratio; CI, confidence intervals.

**Table 3.** Factors associated with overall survival of incident colorectal cancers in National Cancer Database diagnosed during 2010-2014 by tumor deposit status

|  | HR | 95% CI | P |
| --- | --- | --- | --- |
| **Tumor-deposits absent (n=8,110)** |  |  |  |
| MSI (Unstable/High vs Stable/Low) | 1.05 | (0.78 to 1.42) | 0.75 |
| Kras (Mutated vs Wild type) | 1.40 | (1.09 to 1.79) | 0.008 |
| **Tumor-deposits present (n=2,618)** |  |  |  |
| MSI (Unstable/High vs Stable/Low) | 1.65 | (1.11 to 2.47) | 0.014 |
| Kras (Mutated vs Wild type) | 1.63 | (1.16 to 2.28) | 0.004 |
| **Tumor-deposits status not available (n=457)** |  |  |  |
| MSI (Unstable/High vs Stable/Low) | 2.66 | (0.92 to 7.69) | 0.071 |
| Kras (Mutated vs Wild type) | 0.32 | (0.11 to 0.95) | 0.039 |

P of multivariate Cox regression analyses adjusted for age, tumor grade, pathologic stage, Charlson-Deyo score, chemotherapy status, radiotherapy status and race as time-varying covariates; MSI, microsatellite instability.

## Discussion

Here we showed that Kras mutation was more frequent in TDs-positive CRC in a multivariable model. Adjusted to many prognostic and therapeutic factors, Kras mutation was independently linked to a worse overall survival of the CRC with or without TD, but not a better overall survival in the CRC with no known TD status.

The TNM/AJCC 7^th^ and 8^th^ Editions define tumor tissue in peri-colorectal adipose tissue and lymph drainage sites with no histologic evidence of lymph node residues as TD, which possibly indicating discontinuous spread, venous invasion or a totally replaced lymph node.[12, 13] TDs have been reported to be associated with aggressive tumor features, including vascular invasion, perineural invasion, depth of tumor invasion and regional lymph nodes metastasis.[25-28] The RAS family of oncogenes was one of the first to be identified as mutated in human cancer.[29] Ras functions downstream of EGFR and the Kras gene product (Kras protein) is responsible for transduction of mitogenic signals from the EGFR on the cell surface to the cell nucleus. The EGFR signals protect cells from apoptosis, facilitate invasion, and promote angiogenesis and EGFR has been implicated in colorectal tumorigenesis, tumor progression, and metastasis.[30, 31] Considering our finding that Kras mutation was independently associated with tumor-deposit presence and worse survival in CRC, it is possible that Kras mutation also plays a role in the development of tumor deposits and CRC progression. Additional studies of molecular and cell biology are warranted to examine such a role of Kras mutation.

Recently, more and more biological and molecular markers are being used to predict the long-term outcome of CRC patients undergoing surgery. One of the most commonly available biomarkers in the treatment of CRC patients is Kras. Andreyev et al found that the presence of Kras mutation increased the risk of recurrence and death.[2] Maughan et al showed that Kras mutation was a strong negative prognostic factor and the median OS in patients with Kras mutations was significantly shorter than those with wild-type Kras.[9] Some recent studies suggested that no significant prognostic value based on Kras mutation status.[10, 11] However, the prognostic implications of Kras mutations in TDs-positive colorectal carcinoma are still not defined. In our study of a large, national population of patients with CRC, Kras mutation was independently linked to a worse overall survival of the CRCs with or without TDs.

MSI refers to the hypermutable state of cells caused by impaired DNA mismatch repair (MMR). MMR protein status assessment is recommended by the NCCN and European Society for Medical Oncology (ESMO) guidelines for all patients with CRC, especially the patients with resected stage II CRC before adjuvant chemotherapy.[8, 32] A previous study showed deficient

MMR(dMMR)status was associated with improved DFS in the patients treated with surgery alone and no benefit in DFS from fluorouracil (FU)-based treatment was observed for patients with dMMR status.[24, 33] Sinicrope et al revealed that deficient MMR phenotype remains a favorable prognostic factor in patients with stage III colon cancer receiving FOLFOX (folinic acid, fluorouracil, and oxaliplatin) adjuvant chemotherapy and dMMR was significantly associated with better survival after recurrence.[34, 35] In our study, univariate Cox regression analysis and log-rank test both show that there is no association between MSI status and overall survival, which may be attributable to the use of MSI status for choosing chemotherapy. The other reason may be that tumor stage, a critical prognostic factor for CRC, was not included in the univariable analyses.

Several strengths of the study are noteworthy. We used the NCDB, which is a widely used and validated large cancer database.[18] Several clinical prognostic and therapeutic factors were also included in the survival analysis, including Charlson-Deyo Score, radiotherapy status, race and chemotherapy status. Inclusion of these models are expected to reduce, if not eliminate, the potential biases associated with those factors. Moreover, the large sample size of the study lent us the advantages of more statistical power and inclusion of more covariates in analyses. Finally, we included several factors as time varying covariates due to the violation of (constant) proportional hazard assumption. Many studies did not check the violation of (constant) proportional hazard assumption in Cox regression, and may erroneously report the uncorrected HR.

The current study had several limitations. First, as with all retrospective studies, there might be some selection bias despite the adjustment of many covariates. Second, only about 11.5% of the CRC cases in the NCDB had data on Kras mutation. There thus may be a potential selection bias associated with missing data on Kras status. A population-based study is needed to examine our findings. Third, anti-EGFR therapy was associated with Kras status, but not captured in any known population-based cancer databases. Additional validation studies thus are needed. Finally, the interobserver variations in the detection TD, MSI and Kras may exist. Future works using a centralized laboratory are needed to address this issue.

## Conclusions

In this NCDB-based cohort, Kras mutation is independently associated with the tumor deposit presence in CRC, and a worse overall survival in CRC with or without tumor-deposits. Therefore, Kras mutation may play a role in the development of tumor deposits in CRC and serve as a target for CRC treatment, besides guiding anti-EGFR treatment.

## Data Availability

We cannot share the patient-level data which are confidential and were obtained with review and approval from the National Cancer Database and the American College of Surgeons. Aggregated data are available upon request.

## Acknowledgement

The study was in part supported by an Initiative for Multidisciplinary Research Teams (IMRT) award from Rutgers University, Newark, NJ (to N.G. and L.Z.), the U.S. National Institute of Health (R01 AT010243 to N.G.), the National Natural Science Foundation of China (No. 81872387 to M.Z.) and China Scholarship Council (to M.Z.). The funders have no roles in the study design or manuscript preparation.

## Authors’ contributions

MZ, NG, JY and LZ designed the study, LZ extracted the data, MZ, WH, and LZ analyzed the data, MZ and LZ wrote the first draft of the manuscript, and all authors edited and approved the final manuscript.

No conflict of interest is declared by any of the authors.

We cannot share the patient-level data which are confidential and were obtained with review and approval from the National Cancer Database and the American College of Surgeons.

---

**Figure.**
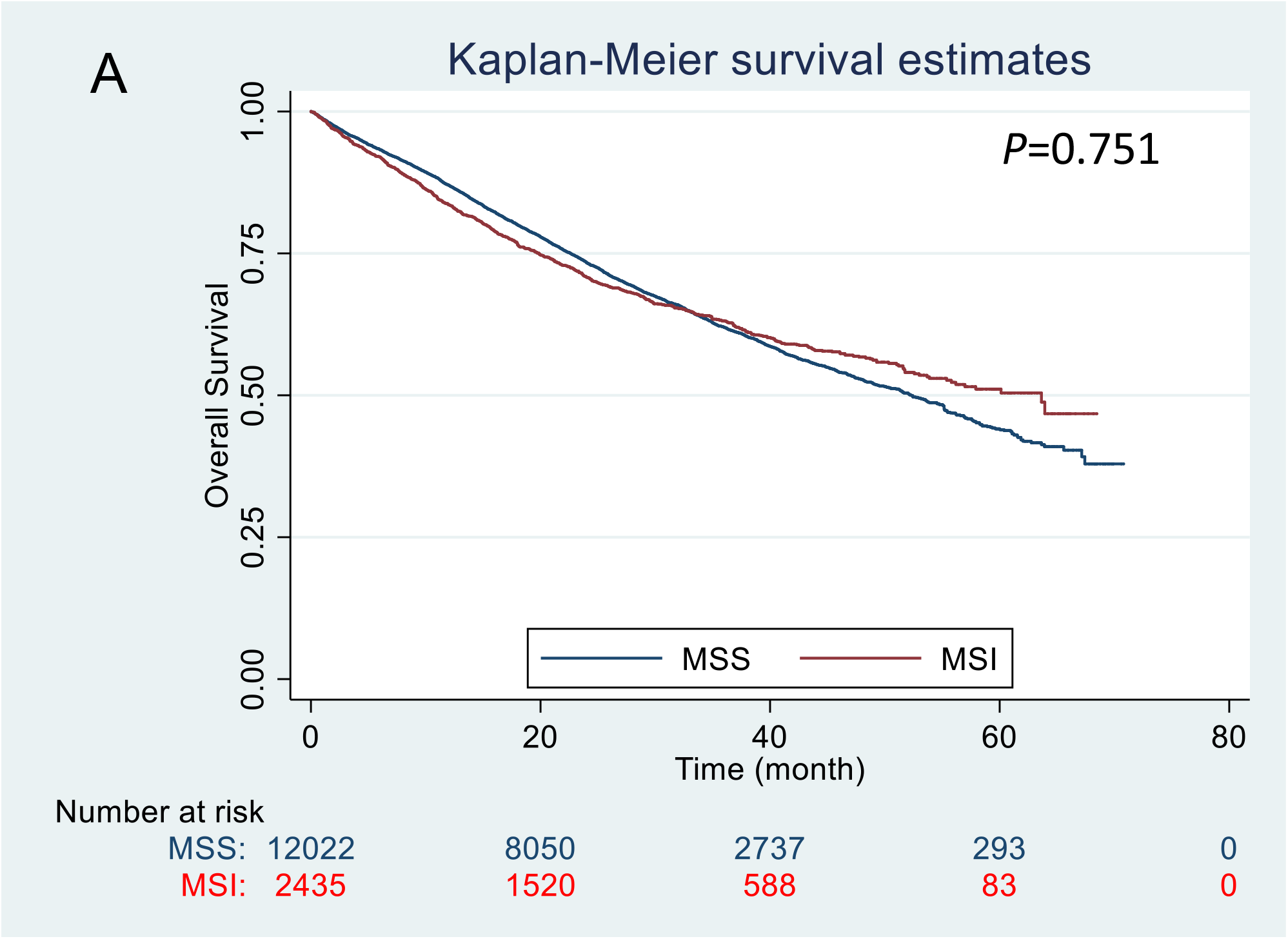
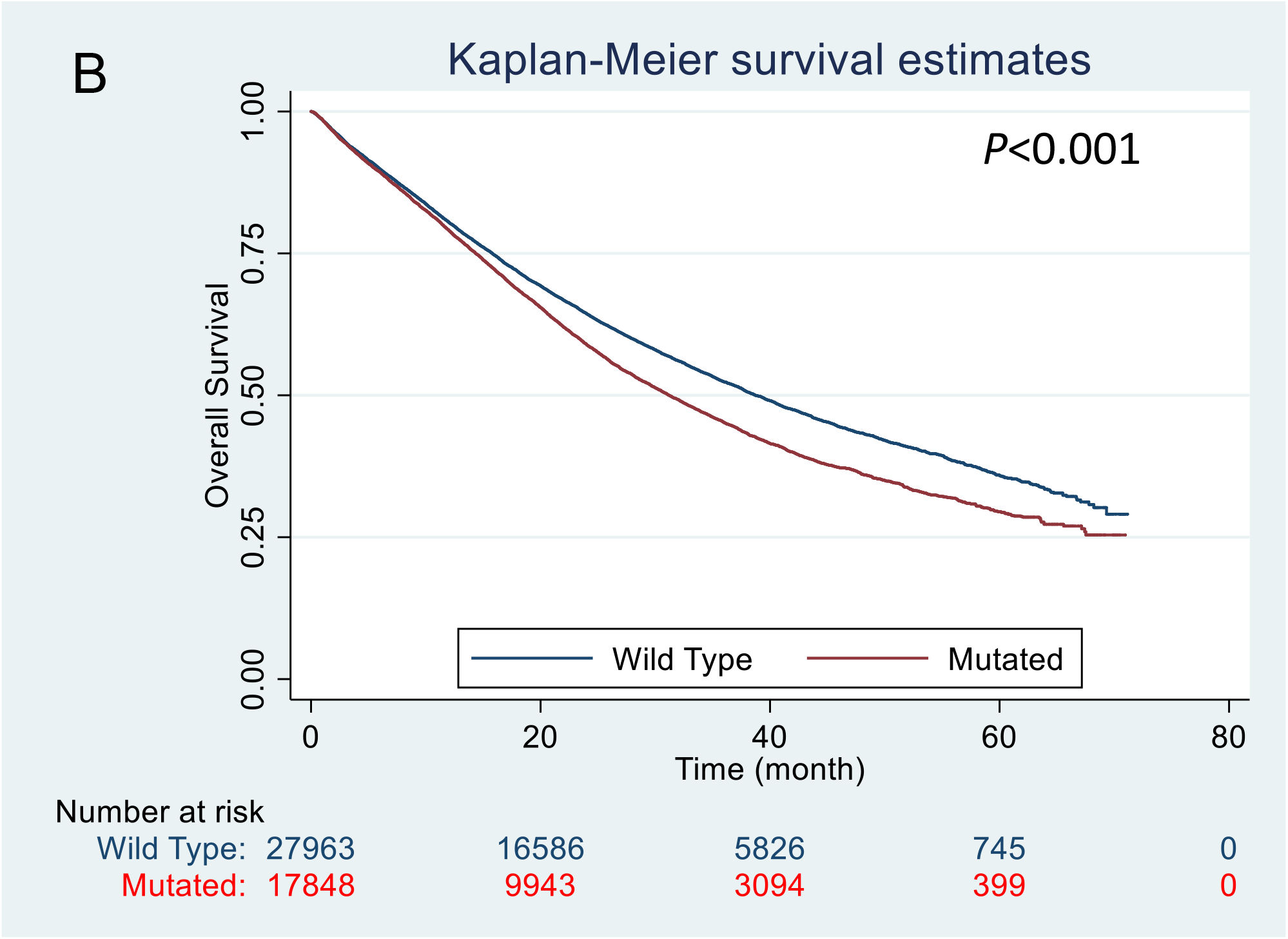
Kaplan-Meier plots of the incident colorectal cancers diagnosed during 2010-2014 in the National Cancer Database that had a known Kras status. **A**. The status of microsatellite instability (MSI) was not associated with overall survival of the colorectal cancers (n=14,457, Log-rank test *P*=0.751). **B**. The Kras mutation (versus wild type) was associated with a worse overall survival of the colorectal cancers (n=45,811, Log-rank test *P*<0.001).

